# Genetic relationships between chronic pain, psychopathologies, and neuroticism

**DOI:** 10.1101/2023.06.20.23291689

**Authors:** Katerina Zorina-Lichtenwalter, Carmen I. Bango, Marta Čeko, Yoni K. Ashar, Matthew C. Keller, Tor D. Wager, Naomi P. Friedman

## Abstract

Chronic pain and psychiatric conditions have consistently demonstrated substantial overlap in risk factors, epidemiology, and effective treatments. Previous work has identified cross-condition latent factors underlying shared genetic risk for several distinct psychiatric conditions and pain conditions. Here, we sought to examine the relationships between these latent genetic factors to determine biological mechanisms common to both pain and psychiatric conditions. We combined two previously published genetic struc-turl equation models. The first model consisted of 24 pain conditions and their two latent factors: General and Musculoskeletal pain-specific. The second model consisted of 11 psychiatric conditions and their four latent factors: Externalizing, Internalizing, Compulsive Thought, and Psychotic Thought. The combined model of six factors and 35 conditions allowed us to estimate correlations between all factors and between conditions of one domain (pain) and factors of the other (psychiatric). We then added three measures of neuroticism (depressive affect subscale, worrying subscale, and total neuroticism score) to this model to examine correlations with all conditions and factors and test for possible explanation of pain-mental disorder relationships by neuroticism. We found that genetic associations between pain and psychiatric conditions were selective to the General Pain factor (and not Musculoskeletal) and Internalizing and Externalizing, but not Thought disorder factors. Neuroticism was associated with pain conditions to the extent that they loaded onto the General Pain factor (i.e., were associated with other pain conditions). Neuroticism also explained a substantial proportion of shared genetic variance between General Pain and Externalizing and between General Pain and Internalizing factors. Overall, the genetic risks shared among chronic pain and psychiatric conditions and neuroticism suggest shared biological mechanisms and underscore the importance of clinical assessment and treatment programs that leverage these commonalities.

## 1 Introduction

Individuals with chronic pain often suffer from multiple forms of psychopathology [84, 126]). Pain and psychiatric conditions correlate at both phenotypic [38, 118, 76] and genetic [77, 58, 41] levels and share psychological [30, 125,91, 69, 116], neurological[32], behavioral [56, 12], and genetic [119] risk factors. Moreoever, chronic pain and psychopathology respond to some of the same drugs [66, 82, 28, 33] and to similar behavioural [49, 98] and psychological [51, 99, 2] interventions. These commonalities suggest shared underlying biological mechanisms that are important to understanding and treating the full extent of the chronic pain experience. Here, we investigate this possible shared biological risk by assessing genetic relationships among chronic pain and related psychosocial constructs.

We recently developed a two-factor genomic structural equation model consisting of a common genetic factor shared by many different pain disorders – which we believe to capture susceptibility to cross-condition chronic pain – and an additional musculoskeletal pain factor [127]. Likewise, a recent publication proposed a model of 4 correlated genetic factors (Externalizing, Internalizing, Compulsive Thought, and Psychotic Thought) [86] explaining the relationships among psychiatric disorders that form several subdomains of general psychopathology [18]. The factors in these models indicate clusters of conditions that may share genetically-driven pathophysiology. As such, they are important steps toward a better understanding of shared biological mechanisms as well as more comprehensive clinical assessment and treatment programs that consider common underlying causes rather than address each condition in isolation. Given the extensively documented comorbidity between pain and psychiatric conditions [117], to advance these goals we sought to examine the genetic correlations between the previously derived factors in their respective domains. Additionally, the shared genetic risk between chronic pain and psychiatric conditions may partially reflect neuroticism, a personality construct that measures the extent to which one experiences the world as distressing and threatening. Neuroticism has been reported to have correlations with specific chronic pain conditions at both genetic [80] and phenotypic levels [46, 22, 6] and to lower pain tolerance [71]. Neuroticism has likewise been shown to be associated with psychiatric conditions [60, 111]. A model that combines neuroticism with the pain-psychiatric genetic factors will enable an assessment of genetic relationships among all three domains and will permit to test whether genetic effects shared between neuroticism and pain or between neuroticism and psychopathology account for any genetic risk shared between pain and psychiatric factors.

To gain a better understanding of the possible biological overlap that both cross-condition and specific chronic pain may have with subjective experience, as measured by different psychopathologies and neuroticism, we attempt to answer the following questions: 1. Which, if any, psychopathologies correlate most strongly with the two pain factors (General and Musculoskeletal)? 2. What is the relationship between neuroticism and each of the genetic factors (two pain and four psychiatric)? 3. Are the relationships between pain and psychiatric factors explained in part by neuroticism sharing genetic risk with both? In elucidating the genetic relationships between pain, psychiatric conditions, and neuroticism, our overarching goal is to characterize the genetic predisposition to chronic pain that operates outside of peripheral injury and tissue characteristics and their attendant pathophysiological mechanisms.

## 2 Methods

### 2.1 Overview

The analyses were as follows: obtain summary statistics on previously done genome-wide association studies (GWASs) for pain and psychiatric conditions and neuroticism scores (full and two subscales); estimate pairwise genetic correlations for all conditions and neuroticism scores; specify and fit a structural equation model (SEM) that combines previously published pain SEM and psychiatric SEM; respecify and fit the combined SEM with the full neuroticism score.

### 2.2 Pain conditions

Genetic association summary statistics for pain conditions were taken from GWASs conducted in the UK Biobank (UKBB) as described previously [127]. Briefly, UKBB study participants, recruited between 2006 and 2010 years and aged 40-69, gave genotypes and were extensively phenotyped using questionnaires, as well as hospital and primary care records. We used individuals identified as White Europeans using genomic principal components, as described in [17]. Sample size varied by condition, ranging from 63,982 to 435,971 people, Table 1. The 24 pain conditions (or conditions with persistent pain as a prominent symptom) came from five UKBB categories: Medical conditions (100074), Health outcomes (713), Self-reported medical conditions (1003), Health and medical history (100036), and First occurrences (1712) were as follows: arthropathies, back pain, chest pain at baseline, chest pain during physical activity, carpal tunnel, chronic widespread pain, cystitis, enthesopathies of the lower limb, enthesopathies elsewhere, gastritis, gout, headache, hip arthrosis, hip pain, irritable bowel syndrome (IBS), knee arthrosis, knee pain, leg pain, migraine, neck/shoulder pain, oesophagitis, rheumatoid arthritis, pain in joint, and stomach pain.

**Table 1:**
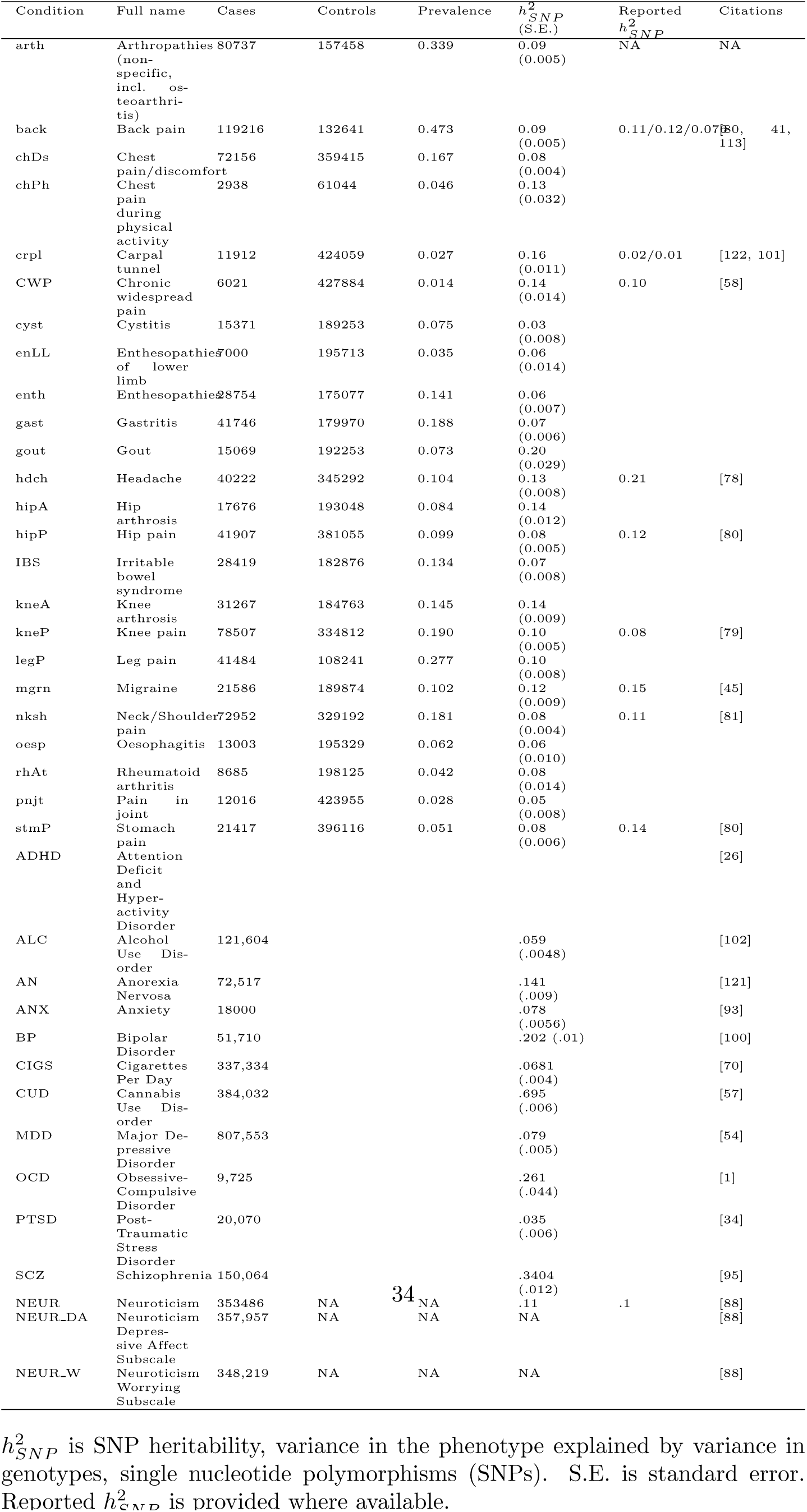
Phenotype descriptive statistics.

### 2.3 Psychiatric conditions

We obtained genetic association summary statistics (effect sizes from regression of the phenotype on single nucleotide polymorphisms from across the genome) for psychiatric conditions from the Psychiatric Genomics Consortium (PGC, https://pgc.unc.edu/for-researchers/download-results/). All studies consisted of a GWAS meta-analysis of several cohorts. If multiple ancestires were included in the original study, we limited our analyses to European White participants. Anxiety (ANX) was a study of 31,060 individuals with measures of five types of anxiety disorders: generalized anxiety disorder, panic disorder, social phobia, agoraphobia, and specific phobias [93]. Major Depressive Disorder (MDD) was a study of 807,553 individuals [54]. Posttraumatic Stress Disorder (PTSD) was a study of 20,730 individuals [34]. Problematic Alcohol Use (ALC) was a study of 141,932 individuals [102], using scores from the Alcohol Use Disorders Identification Test (AUDIT) [104]. Cigarettes per Day (CIGS) was a study of 337,334 individuals [70]. Cannabis Use Disorder (CUD) was a study of 374,287 individuals [57]. Attention-Deficit/Hyperactivity Disorder (ADHD) was a study of 55,374 individuals [26]. Schizophrenia (SCZ) [95] was a study of 150,064 individuals. Bipolar Disorder (BP) [100] was a study of 74,194 individuals. Obsessive Compulsive Disorder (OCD) was study of 9,725 individuals [1]. Anorexia Nervosa (AN) was a study of 72,517 individuals [121].

### 2.4 Neuroticism

We obtained genetic association summary statistics for neuroticism (https://ctg.cncr.nl/software/summary_statistics) [88], which came from the meta-analysis of a GWAS Nagel and colleagues conducted in the UKBB and another meta-analysis conducted previously by the Genetic Personality Consortium (GPC) [24]. In the UKBB, the neuroticism score was a count of ”Yes” entries in response to the following 12 questions: 1. ”Does your mood often go up and down?” 2. ”Do you ever feel ’just miserable?” 3. ”Are you an irritable person?” 4. ”Are your feelings easily hurt?” 5. ”Do you often feel ’fed-up’?” 6. ”Would you call yourself a nervous person?” 7. ”Are you a worrier?” 8. ”Would you call yourself tense or ’highly strung’?” 9. ”Do you worry too long after an embarrassing experience?” 10. ”Do you suffer from ’nerves’?” 11. ”Do you often feel lonely?” 12. ”Are you often troubled by feelings of guilt?”. The sample size for this meta-analysis was 389,581. Additionally, we used summary statistics for GWASs of two subscales of the UKBB neuroticism scale, which measured the dimensions depressive affect (questions 1, 2, 5, and 11) and worrying (questions 6, 7, 8, and 10) [89]. The sample sizes for these dimensions were 311,866 for depressive affect and 305,202 for worrying.

### 2.5 Heritability and genetic correlations

Using the Genomic SEM R package, we converted summary association statistics from GWAS for all phenotypes to z-scores with the **munge** function, and we estimated genetic covariances and single nucleotide polymorphism (SNP) heritabilities with the **ldsc** function [15], as implemented in the Genomic SEM R package, described in section 2.6. SNP heritability (*h*^2^) is the variance explained in the phenotype by the variance in the SNP allele counts. For binary (yes/no) conditions, we used effective sample size (provided with the summary statistics or calculated using the method described in [47]) to estimate genetic variance and heritability. We converted genetic covariances to correlations using the **cov2cor** function in R, which produced a matrix of genetic correlations across all pairs of conditions and neuroticism scores. A genetic correlation (*r_g_*) estimates the relationship (direction and magnitude) between the variant-allele effects on one condition and the variant-allele effects on the other, using all available genetic variants. It thus shows the extent to which two conditions or traits share genetic predisposition. Pairwise genetic correlation estimates are a function of the product of both conditions’ SNP heritabilities:

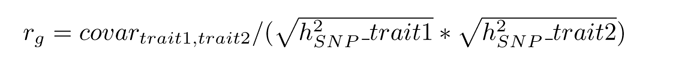

We also calculated the Pearson correlation between pain condition loadings onto the General Pain factor and the correlations between all pain conditions and neuroticism. For this estimation, we used the **corr** function in R.

### 2.6 Structural equation modelling

To estimate the magnitude and direction of correlation between previously derived latent genetic factors, we used structural equation modeling (SEM), specifically as implemented for use with genetic scores in Genomic SEM [48]. The SEM framework is intended for extracting shared variance among several indicators (here, pain and psychiatric conditions) with latent factors. The relationships between the indicators and their latent factors are specified by the researcher with a structural model (such as in Figure 1a) and 1b) based on a theoretically or empirically driven hypothesis. In the structural model, the arrows and their associated numbers (factor loadings) correspond to coefficients from linear regression, simultaneously run for all indicators specified for a given latent factor. In their standardized form, these loadings represent the square root of the proportion of variance explained in the given indicator by the latent factor. In addition to estimating factor loadings, it is possible to estimate inter-factor correlations, which is the main goal of this analysis for the present study. We used a 24-condition two-factor pain model [127] and an 11-condition four-factor psychiatric model [86] to specify a combined model with all six factors allowed to correlate. Given that Genomic SEM takes as input a matrix of genetic correlations, we estimated all pairwise genetic correlations for the complete set of 35 conditions (plus three neuroticism scores for subsequent analyses) in LDSC (Section 2.5). As done for the original constituent models, we specified all 24 pain conditions to load onto the General Pain factor, and 11 of these conditions to load onto the Musculoskeletal factor, [127]. For the psychiatric portion, we specified ADHD, ALC, CIGS, and CUD to load onto the Externalizing factor, SCZ and BP onto the Psychotic Thought factor, OCD and AN onto the Compulsive Thought factor, and MDD, ANX, and PTSD onto the Internalizing factor [86]. Given that the model as described above did not have optimal fit, we examined residual covariances for outliers, i.e. pairwise covariances between conditions that were not adequately explained by the model. Among the particularly large residual covariances, several had previously reported comorbidities: CIGS with chest pain [120], stomach pain with psychotic disorders [123, 29] and AN [50] and IBS with both COM factor conditions: OCD [115] and AN [75]. Therefore, we further specified in the model and estimated residual covariances for the following condition and condition-factor pairs: CIGS with chest pain (both at baseline and during physical activity); stomach pain with SCZ and AN; and IBS with the Compulsive factor.

**Figure 1:**
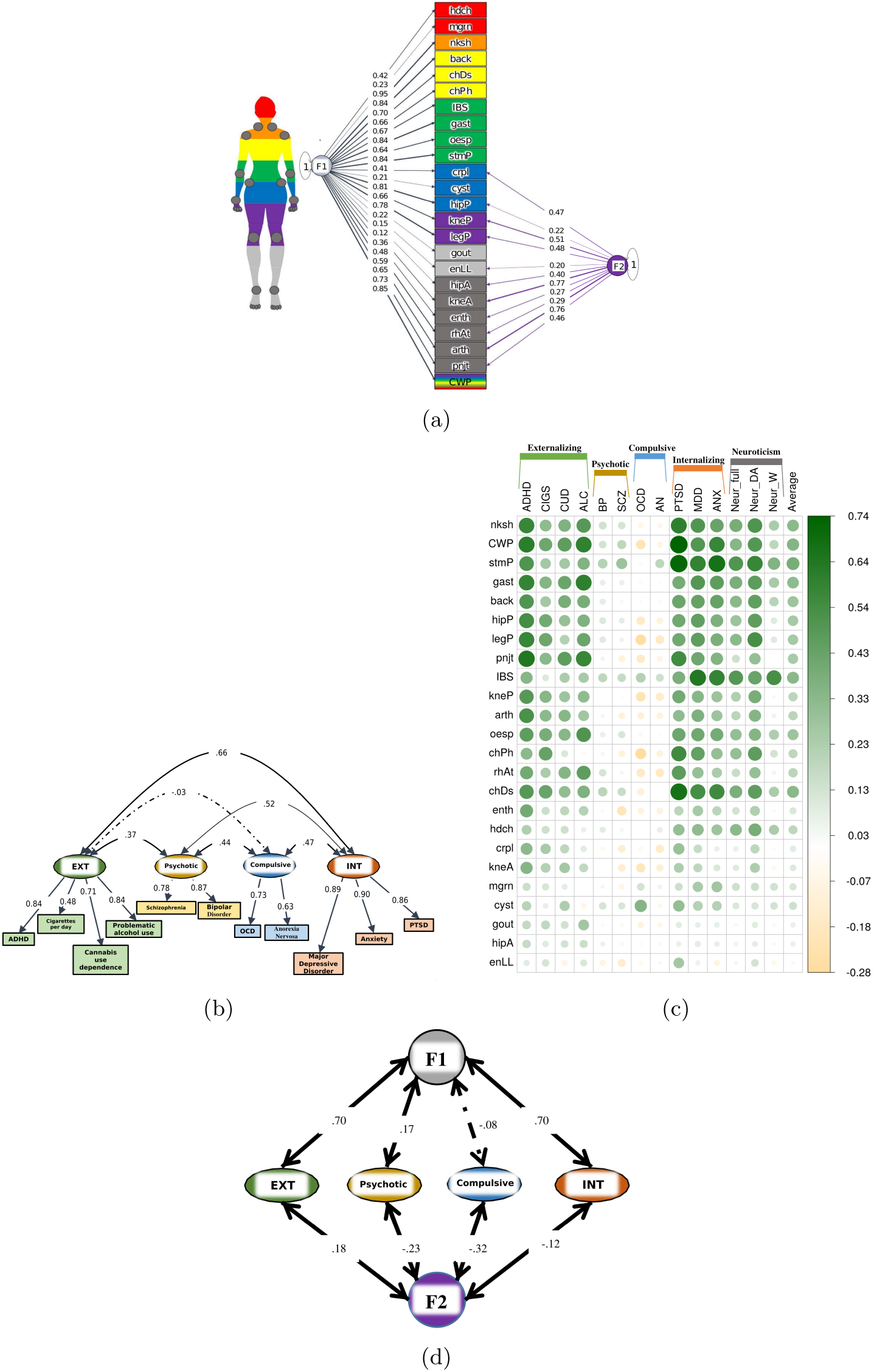
Pain and psychological models and correlations. (A) Genomic SEM model of 24 pain conditions from [127], with all conditions loading onto the General Pain factor, F1, and 11 conditions loading onto the Musculoskeletal pain factor, F2 (see **Section 3.2**. Loadings are shown on the arrows. Additional residual correlations were estimated for the following same body-site pairs (not shown in figure): chest pain at baseline and during physical activity; hip arthrosis and hip pain; knee arthrosis and knee pain, migraine and headache. Comparative fit index (CFI) = .956; Standardized Root Mean Square Residual (SRMR) = .075. (B) Genomic SEM model of 11 psychiatric conditions from [86], with four factors: Externalizing (EXT), Psychotic, Compulsive, Internalizing (INT). CFI = .960; SRMR = .082. (C) Genetic correlations estimated in LDSC (linkage disequilibrium score regression) for pain conditions with psychiatric conditions and neuroticism. The pain conditions are listed in descending order of their loading onto F1, and psychiatric disorders are grouped by their factors and listed in descending order of their loading onto F1 within each psychiatric factor, left to right. The average cross-column correlations with psychiatric conditions and neuroticism scores are given in the ”Average” column. (D) Combined Genomic SEM model of pain and psychiatric conditions with inter-factor correlations shown on the arrows. Correlations between psychiatric factors were similar to those in B. and are left out for clarity. In addition to the residual correlations specified for the pain SEM (A.), the following residual correlations were estimated (also not shown in figure): CIGS with chest pain at baseline and during physical activity; stomach pain with SCZ and AN; IBS with the Compulsive factor. CFI = .907; SRMR = .075. Pain conditions: nksh, neck/shoulder pain; CWP, chronic widespread pain; stmP, stomach pain; gast, gastritis; back, back pain; hipP, hip pain; legP, leg pain; pnjt, pain in joint; IBS, inflammatory bowel syndrome; kneP, knee pain; arth, arthritis; oesp, oesophagitis; chPh, chest pain during physical pain; rhAt, rheumatoid arthritis; chDs, chest pain; enth, enthesopathies; hdch, headache; crpl, carpal tunnel syndrome; kneA, knee arthrosis; mgrn, migraine; cyst, cystitis; gout, gout; hipA, hip arthrosis; enLL, enthesopathies of lower limb. Psychiatric conditions: ADHD, attention deficit and hyperactivity disorder; CIGS, cigarettes per day; CUD, cannabis use disorder; ALC, alcohol use disorder; BP, bipolar disorder; SCZ, schizophrenia; OCD, obsessive compulsive disorder; anorexia nervosa; PTSD, posttraumatic stress disorder; MDD, major depression disorder; ANX, anxiety. Neuroticism scores: Neur full, full scale; Neur DA, depressive affect subscale of neuroticism; Neur W, worry subscale.

We evaluated the models using CFI (comparative fit index), which compares the model fit to one in which the variables are entirely independent, and SRMR (standardized root mean squared residual), a measure of variance unexplained by the model [55]. A well-fitting model should generally have a CFI*≥ .*95 and an SRMR*≤ .*08 [55], although lower CFI and higher SRMR thresholds are acceptable in the context of Genomic SEM [48].

Because of the high correlation between Externalizing and Internalizing, we additionally estimated correlations between the General Pain factor and Internalizing and Externalizing factors, each psychiatric factor independent of the other psychiatric factor. We used Cholesky decomposition [63], a method that decomposes a symmetric matrix, such as the covariance matrix of our genetic factors, and provides estimates of shared and independent variance components. We thus specified two models with the following factors: Externalizing and Internalizing, two higher-order factors we called X1 and X2, and the two pain factors. The higher-order factors represent the following, respectively: variance in the first psychiatric factor that is also allowed to predict the second psychiatric factor and residual variance in the second psychiatric factor that is independent of the first. The two models allowed us to examine the semipartial correlations of the pain factors with the psychiatric factor predicted by X2 (i.e., partialling out the other psychiatric factor). Thus, the key estimates of interest are the X2 factor correlations with the two pain factors, General Pain (F1) and Musculoskeletal Pain (F2), because they provide estimates of correlations between pain factors and the unique variance in each psychiatric factor.

We additionally estimated correlations between the pain factors and each psychiatric condition, between the psychiatric factors and each pain condition, and between neuroticism and each condition from both sets. For common cause analyses, we estimated genetic relationships between neuroticism, the General Pain factor and psychiatric factors. Namely, for each psychiatric factor, we specified two bidirectional paths between this factor and the General Pain factor: one direct path and one indirect path, which goes through neuroticism. We calculated the proportion of the correlation between the pain and psychiatric factors that was due to neuroticism by dividing the indirect path coefficient (product of the path coefficients between neuroticism and each factor) by the total genetic relationship between General Pain and the psychiatric factor (direct path coefficient + indirect path coefficient).

## 3 Results

### 3.1 Pain-Psychiatric Correlations

We combined 24 pain conditions from our previously published pain model, Figure 1a [127], 11 psychiatric conditions from a published psychiatric model (Figure 1b [86]), and neuroticism (full scale and two subscales), and we calculated their pairwise genetic correlations. In the correlation matrix (Figure 1c) the pain conditions are ordered by loading onto the General Pain factor (F1 in Figure 1a), descending from top to bottom. We observe that the pain conditions in the top 75% of the correlation matrix (which have the top 75% of the General Pain factor loadings), from neck/shoulder pain to knee arthrosis, have the largest genetic correlations with a number of psychiatric conditions and with two neuroticism scales.

Interestingly, the pain-psychiatric condition correlations show patterns segregating by psychiatric factor (Figure 1b): Internalizing (Section 3.1.1) and Externalizing (Section 3.1.2) factors show positive and substantial correlations with pain conditions, while the Psychotic factor (Section 3.1.3) shows weak cor-relations, and the Compulsive (Section 3.1.4) factor shows weak and mostly negative correlations. The pain condition-neuroticism correlations are strongest for the full neuroticism scale and the depressive affect sub-scale (see Section 3.1.5). We discuss these correlations (grouped by psychiatric factor) and neuroticism below.

#### 3.1.1 Internalizing Conditions with Pain Conditions

The psychiatric factor with the strongest genetic relationships with pain is Internalizing, consisting of MDD, ANX, and PTSD. Approximately 75% of the pain conditions have genetic correlations *> .*2 with each Internalizing condition: 19 with MDD, 18 with ANX, and 20 with PTSD. The pain conditions correlated with all three Internalizing conditions fall into the following categories: cranial (headache and migraine), visceral (gastritis, oesophagitis, chest pain, IBS, and stomach pain), and musculoskeletal (arthrosis, back, neck, hip, knee, leg, joint, rheumatoid arthritis (RA) and chronic widespread pain (CWP, which is a measure of pain all over the body, corresponding to the UK Biobank datafield 2956 and is distinct from multi-site pain, defined as a count of pain sites across the body and reported in [58, 65]). PTSD is the only psychiatric condition with a *> .*7 genetic correlation with pain: CWP and stomach pain (.74 and .72, respectively).

#### 3.1.2 Externalizing Conditions with Pain Conditions

Externalizing is the factor with the next most numerous and highest correlations with pain, for all four of its indicators (CIGS, CUD, ADHD, ALC), (Figures 1 and 2a). Approximately 75% of the pain conditions have genetic correlations *> .*2 with each Externalizing condition: 20 with ADHD, 16 with CIGS, and 16 with CUD. The pain conditions correlated with all four Externalizing conditions are musculoskeletal: neck/shoulder, CWP, back, hip, leg, joint pain, knee, and arthrosis; and visceral: chest, stomach, gastritis, and oesophagitis.

**Figure 2:**
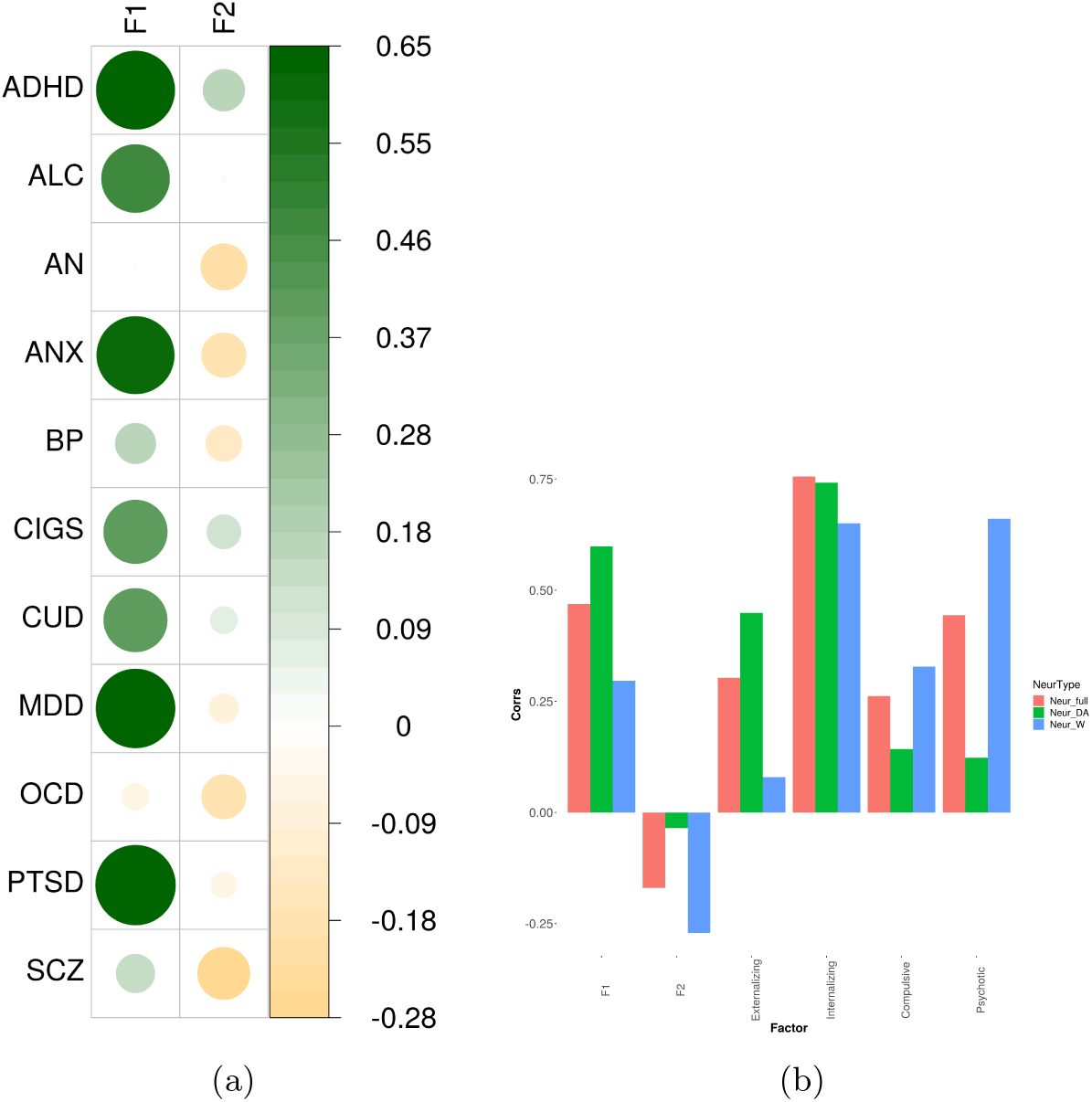
(A) Factor-condition correlations for psychiatric conditions with pain factors. (B) Correlations between neuroticism and all factors. Pain conditions: nksh, neck/shoulder pain; CWP, chronic widespread pain; stmP, stomach pain; gast, gastritis; back, back pain; hipP, hip pain; legP, leg pain; pnjt, pain in joint; IBS, inflammatory bowel syndrome; kneP, knee pain; arth, arthritis; oesp, oesophagitis; chPh, chest pain during physical pain; rhAt, rheumatoid arthritis; chDs, chest pain; enth, enthesopathies; hdch, headache; crpl, carpal tunnel syndrome; kneA, knee arthrosis; mgrn, migraine; cyst, cystitis; gout, gout; hipA, hip arthrosis; enLL, enthesopathies of lower limb. Psychiatric conditions: ADHD, attention deficit and hyperactivity disorder; CIGS, cigarettes per day; CUD, cannabis use disorder; ALC, alcohol use disorder; BP, bipolar disorder; SCZ, schizophrenia; OCD, obsessive compulsive disorder; anorexia nervosa; PTSD, posttraumatic stress disorder; MDD, major depression disorder; ANX, anxiety; F1, General Pain factor; F2, Musculoskeletal pain factors. Neuroticism scores: Neur full, full scale; Neur DA, depressive affect subscale of neuroticism; Neur W, worry subscale.

#### 3.1.3 Psychotic Thought Conditions with Pain Conditions

SCZ has a positive correlation *> .*2 with stomach pain (.28) and a negative correlation (*−.*2) with enthesopathies (*−.*21). BP also has a positive correlation *> .*2 only with stomach pain.

#### 3.1.4 Compulsive Thought Conditions with Pain Conditions

OCD’s substantial associations with pain are limited to cystitis (.35) and IBS (.22) and negative correlations with 4 conditions (*< −.*2), (Figures 1 and 2a): CWP, chest pain during physical activity, knee pain, and leg pain. AN has no genetic correlations with pain *> .*2 or *< −.*2.

#### 3.1.5 Neuroticism with General Pain

We hypothesized that the magnitude of loading for a given pain condition onto the General Pain factor (F1 in Figure 1a) may be related to that pain condition’s magnitude of correlation with neuroticism, Figure 1c. To test this, we examined the pattern of these loadings and compared it to correlations with neuroticism across pain conditions. We found the loading pattern to be similar to the pattern of correlations with the full neuroticism scale as well as the depressive affect subscale. To quantify the similarity in these patterns, we estimated the Pearson correlation between the factor loadings and the pain-neuroticism correlations and found it to be .80 for the full-scale, .93 for the depressive affect subscale, and .51 for the worry subscale.

### 3.2 Pain-Psychiatric SEM

The SEM fit well, CFI = .907 and SRMR = .075. The inter-factor correlations (Figure 1d) are the strongest for the General Pain Factor with Externalizing and Internalizing (both .70). The correlation with the Psychotic factor is .17 and not significant with the Compulsive factor. For the Musculoskeletal factor, there is a weak correlation with Externalizing (.18) and negative correlations with the remaining psychiatric factors (*−.*23*, −.*32, and *−.*12).

The additional estimated path coefficients for residual covariances for stomach pain with SCZ and AN, IBS with the COM factor, and chest pain with CIGS improved the fit (without these paths, CFI = .883; SRMR = .078). Additionally, adding these estimates did not substantially alter the factor loadings and inter-factor covariances, the biggest difference for which was .03 (in standardized space).

The high correlations for the General Pain Factor with both Externalizing and Internalizing, which are themselves highly correlated (.67 (Figure 1b), led us to examine the correlation for each of these psychiatric factors with the pain factor independent of the other (i.e. Externalizing independent of Internalizing and vice versa) (Figure 3). The correlation between Internalizing and General Pain independent of Externalizing is .30 and the correlation between Externalizing and General Pain independent of Internalizing is .33.

**Figure 3:**
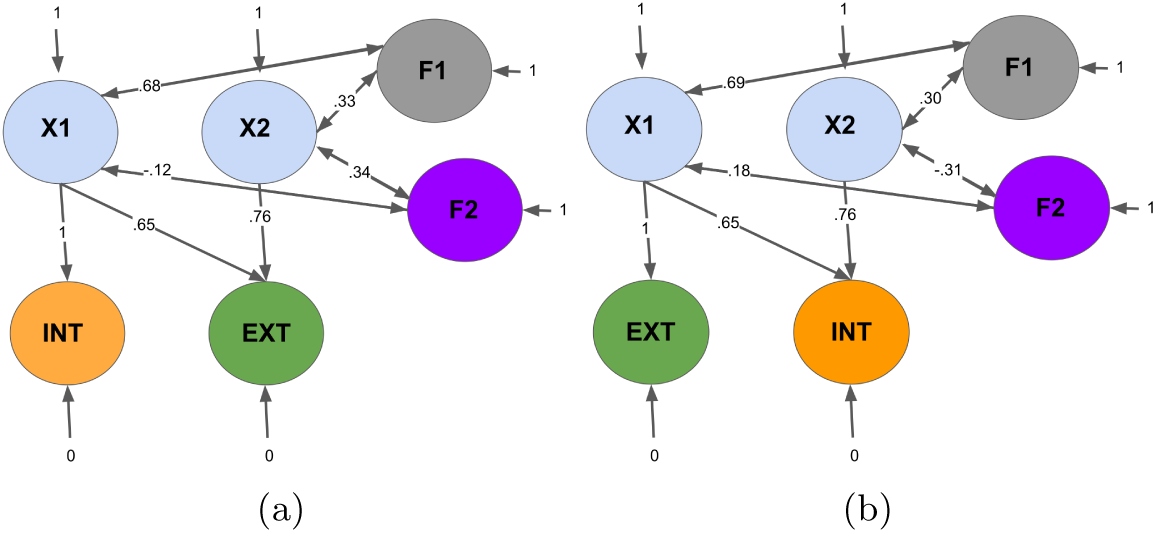
Cholesky decomposition. EXT, INT, and F1, are lower-order latent factors – Externalizing, Internalizing, and General Pain, defined as shown in Figure 1. X1-X2 are arbitrarily named higher-order latent latent constructs that capture shared and independent variance in Externalizing and Internalizing. In subfigure (A), The numbers on the arrows from X1 and X2 to INT and EXT represent the following: 1 is the standardized variance in Internalizing that is also allowed to predict Externalizing; .65 is the square root of the standardized covariance in Externalizing with Internalizing; .76 is the square root of the standardized variance in Externalizing independent of Internalizing. The double arrows between F1, F2 and X1 represent standardized covariance between General and Musculoskeletal Pain, respectively, and the standardized variance of Externalizing that includes its covariance with Internalizing. The double arrows between F1, F2, and X2 represent the standardized shared variance between General and Musculoskeletal Pain, respectively, and Externalizing, independent of Internalizing. In subfigure (B), the numbers on the arrows from X1 and X2 to EXT and INT represent the following: 1 is the standardized variance in Externalizing that is also allowed to predict Internalizing; .65 is the square root of the standardized covariance in Internalizing with Externalizing; .76 is the square root of the standardized covariance in Internalizing independent of Externalizing. The double arrows between F1, F2 and X1 represent standardized covariance between General and Musculoskeletal Pain, respectively, and the standardized variance of Internalizing that includes its covariance with Externalizing. The double arrows between F1, F2, and X2 represent the standardized shared variance between General and Musculoskeletal Pain, respectively, and Internalizing, independent of Externalizing. In both subfigures, the double arrows between F1, F2, and X2 provide the answer to the question that instigated this analysis: there is shared variance between both General Pain and Musculoskeletal Pain and Externalizing, independent of Internalizing (.33 and .34, respectively, standardized); and there is shared variance between both General Pain and Musculoskeletal Pain and Internalizing, independent of Externalizing (.30 and *−.*31, respectively, standardized). For both models, the comparative fit index (CFI) was .917, the Standard3i1zed Root Mean Squared Residual (SRMR) was .074, and chi-square was 20440, with 523 degrees of freedom.

### 3.3 Pain-Psychiatric condition-factor correlations

Next we wanted to ensure that the correlations between pain and psychiatric factors (shown in Figure 1d), accurately reflect the composition of each factor, i.e., that the correlation between General Pain and Externalizing is not driven by CIGS, for example, despite the fact that CIGS is the weakest indicator of Externalizing. Thus, we estimated the correlations between pain factors and psychiatric conditions (Figure 2). The correlations for the four psychiatric conditions of the Externalizing factor are strong for General Pain (.41 *− .*62), with ADHD being the strongest. This is consistent with the loadings of these four disorders onto their factor, Externalizing, such that CIGS and CUD are moderate (.48 and .71, respectively) and ALC and ADHD are high (both .84), Figure 1b. All three Internalizing factor conditions MDD, ANX, and PTSD have strong correlations with General pain (.61 *− .*65), consistent with the psychiatric condition loadings on the Internalizing factor (.86 *− .*90). For the thought factors, all four of the conditions have weak and/or negative correlations with General Pain. The correlations between the Musculoskeletal factor and psychiatric conditions are low for two of the Externalizing conditions (ADHD, .18, CIGS, .12) not significant for the other two Externalizing conditions (CUD, ALC) and PTSD, and negative for both Thought factor disorders and remaining Internalizing factor disorders (SCZ, BP, OCD, AN, MDD, and ANX). These estimates are consistent with the inter-factor correlations for the Musculoskeletal factor, which are weak (.18, Externalizing) and weak negative (both Thought and Internalizing factors), Figure 1d. Thus, the inter-factor correlations (Figure 1d) accurately represent the composition of the psychiatric factors (Figure 1b).

### 3.4 Neuroticism correlations with pain conditions

A previously published phenotypic factor analysis revealed three factors extracted from the neuroticism subscale in the Eysenck Personality Questionnaire, Revised in 1985 (EPQ-R; [40]), two of which were depressive affect and worrying [85]. Consistent with these findings, a recent study found the 12-question UKBB neuroticism scale to have two genetic subclusters: the first characterized by loneliness, miserableness, mood swings, and ”fed-up” feelings; the second characterized by nervousness, worrying, tenseness, and suffering from ”nerves” [89]. Along with the full neuroticism scale score GWAS summary statistics, we downloaded the GWAS summary statistics for each subcluster (depressive affect and worrying) and estimated the genetic correlations between all three neuroticism scores and our pain conditions (Figure 1c). These estimates show strong correlations for a large number of musculoskeletal and visceral pain conditions as well as headache with depressive affect. For worrying, there are low-moderate correlations (*> .*2 and *< .*4) for neck/shoulder, stomach pain, gastritis, oesophagitis, chest pain, headache, and IBS, with IBS being the strongest by far (.53). Thus the depressive affect dimension of neuroticism as measured in the UKBB shares its genetic risk with many more of the queried pain conditions than the worrying dimension.

### 3.5 Neuroticism as an explanation for shared pain-psychiatric variance

We estimated genetic correlations for neuroticism and its two subscales with each factor (Figure 2b) and then estimated the contribution of neuroticism to the correlations between pain and psychiatric factors (Figure 4). For General Pain and Internalizing, neuroticism explains 39% of the shared variance; for General Pain and Externalizing, neuroticism explains 16% of their shared variance; for General Pain and Psychotic, neuroticism explains 83% of the shared variance (which was already small, Figure 1d); the correlation between General Pain and Compulsive is not significant, Figure 1d.

**Figure 4:**
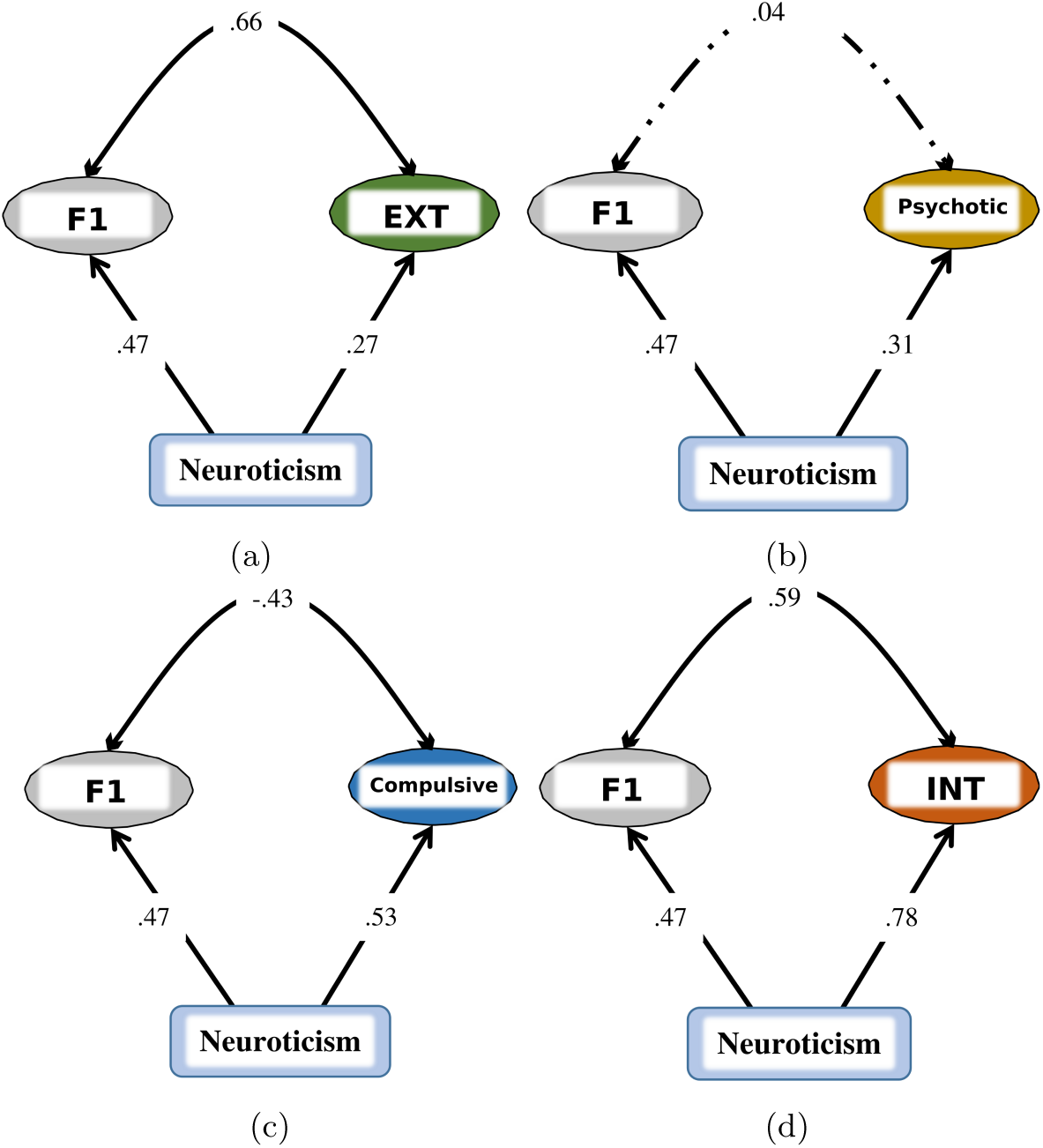
Genomic SEM models of neuroticism as a common cause. Each panel, (A)-(D), shows the correlations for neuroticism with F1, the General Pain factor, and one of the Psychiatric factors (EXT, externalizing, INT, internalizing). For General Pain and INT, neuroticism explains 39% of the shared variance; for General Pain and EXT, neuroticism explains 18% of their shared variance; for General Pain and the Psychotic factor, neuroticism explains 91% of the shared variance; for General Pain and the Compulsive factor, neuroticism explains 19% of their shared variance and reverses the direction of their negative correlation to positive. F2, the Musculoskeletal pain factor was included in the model but is not shown in the figure. The comparative fit index (CFI) = .811, Standardized Root Mean Squared Residual (SRMR) = .077, and chi-square was 644143, with 552 degrees of freedom.

## 4 Discussion

We report a systematic study of genetic correlations between 24 chronic pain conditions in a 2-factor model [127] and 11 psychiatric conditions in a 4-factor model [86], both at the individual condition and factor levels. This analysis enabled us to observe trans-diagnostic genetic risks that bring attention to shared biological mechanisms for conditions previously held to be distinct and treated as such in the clinical setting. Our assessment of shared genetic variance for these two domains, in turn, allowed for a more accurate conceptualization of conditions and disorders whose diagnoses and treatments are frequently obfuscated and complicated by comorbidity [13, 44] and overlapping symptoms [83, 96, 7, 16].

We found that internalizing and externalizing disorders as well as neuroticism and the depressive affect subscale of neuroticism share a considerable amount of genetic risk with chronic pain conditions. At the level of cross-condition factors, internalizing and externalizing disorders likewise share genetic risk with cross-condition chronic pain. Further, the pain conditions that most correlate with neuroticism and its depressive affect subscale share the most genetic variance with other pain conditions. Lastly, the genetic overlap between cross-condition pain and both externalizing and internalizing disorders may be, in part, due to the shared genetic risk among all of these conditions and neuroticism.

These findings provide evidence at the genetic level for shared biological risks for chronic pain above and beyond direct stimulation of primary nociceptors by tissue damage. These supra-nociceptive risks are consistent with the biopsychosocial model of pain, or a dynamic interaction between psychological, social, and pathophysiological factors [90, 43, 9, 11]. In particular, our study offers evidence for a common genetic predisposition that cuts across psychological disorders, the neuroticism dimension of personality, and pain conditions. This common predisposition suggests that in addition to epidemiological interactions between these factors, a single genetic component may underlie their comorbidity. The general pathology corresponding to this genetic predisposition lies at the intersection of, at least, cross-condition chronic pain, externalizing and internalizing psychiatric disorders, and neuroticism (specifically its depressive affect subscale). Though chronic pain is often conceptualized and treated as a primarily biomedical condition driven by peripheral tissue pathology, our results indicate shared mechanisms with psychiatric conditions driven by neurobiological and psychological processes.

Internalizing disorders show the highest genetic correlation with the cross-condition chronic pain factor, which is consistent with previous reports [80, 58, 19]. It has likewise been previously shown that the internalizing disorders we considered here have high comorbidity with chronic pain. Up to 33% of people with depression report chronic pain, and up to 75% of people with chronic pain report depression [124]. Anxiety affects up to 60% of chronic pain patients [53]. PTSD is also highly comorbid with chronic pain [8, 4, 106]. Prior evidence suggests a bidirectional causal relationship between pain and psychiatric conditions: pain is inherently depressing and limits behavior, and depression predisposes individuals to developing chronic pain. Our results advance our understanding of this relationship by indicating a substantial overlap in genetic risk factors that predisposes individuals to both of these conditions.

All four of the externalizing factor conditions (three substance use and ADHD) have substantial correlations with many pain conditions. Substance use disorders in chronic pain patients – including cannabis, alcohol, and cigarettes – have recently been reviewed by Martel et al. [74]. All three disorders, as defined by the Diagnostic and Statistical Manual of Mental Disorders, version 5 (DSM-5), have a higher prevalence in chronic pain patients. CUD is more common among people with chronic pain who also have depression [35], anxiety [25], or other mental health problems [108]. Cigarette smoking has a bidirectional relationship with chronic pain: pain leads to increased use of cigarettes [31], and cigarette smoking leads to chronic pain [103, 105, 107]. Similarly, alcohol use disorder is more prevalent in chronic pain patients [72, 37, 27], and heavy alcohol use may lead to the development of chronic pain [20, 68, 21]. Anxiety [36, 67] and depression [36, 52] as chronic pain comorbidities have shown higher association with heavy alcohol use as well. ADHD has been found to be comorbid with chronic pain in women [5] and increased pain sensitivity has been reported in adults with ADHD [114]. Neuroinflammation may be a linking mechanism [64], and there is evidence that ADHD is under-diagnosed in chronic pain patients [62].

Psychotic (SCZ and BP) and compulsive (OCD and AN) thought disorders have almost no positive genetic correlations with chronic pain conditions, which suggests a lack of shared biological susceptibility. On the other hand, bipolar disorder has been reported to be highly comorbid with chronic pain [109, 10, 92]. There is evidence from two studies that it is genetically correlated with multisite pain [59, 19], which is defined in both studies as the count of chronic pain sites across the body, using self-report in the UKBB. We used the same dataset and questionnaire, but we did not construct a phenotype using the count of pain sites, because as a measure of pain widespreadness it was of limited utility for determining shared cross-condition genetic risk (see [127] for a detailed discussion of our reasons). Our closest phenotype, chronic widespread pain (CWP) shows a genetic correlation of .13 with bipolar disorder. Schizophrenia has been reported to be anti-correlated with chronic pain [94], and people with this diagnosis have a lower intensity of pain compared to other psychiatric disorders and compared to healthy controls [39]. This seemingly protective effect may be due to schizophrenia patients under-reporting chronic pain [110]. There is also evidence for a genetic correlation between schizophrenia and chronic multi-site pain (or count of pain sites, as above) [19].

Of the compulsive thought disorders, anorexia has been reported to be comorbid with abdominal pain [97] and migraine [87] but not to have genetic correlations with chronic pain [19]. There is evidence that OCD is comorbid with chronic pain from population-based but not clinic-based studies [23]. To our knowledge, genetic correlations between OCD and chronic pain have not been assessed previously.

We found that many chronic pain conditions are genetically correlated with neuroticism, particularly the depressive affect subscale, which captures loneliness, miserableness, and moodiness. The pain conditions that had the most shared variance with other pain conditions were more strongly genetically correlated with neuroticism. Several studies have shown phenotypic correlations between neuroticism and different chronic pain conditions. For example, neuroticism is highly prevalent in migraine patients [3], especially in the presence of depression and anxiety [42], as well as neck, shoulder, and back pain [14], and fibromyalgia [73]. One study showed that in anticipation of pain there was a negative correlation between neuroticism and brain activity in emotional and cognitive pain processing areas (parahippocampus, insula, thalamus, and anterior cingulate cortex) [22]. Neuroticism is likewise associated with greater pain catastrophizing (tendency toward exaggerated negative reaction to pain [112]) and pain-related anxiety [61]. Two studies have reported genetic correlations for neuroticism with chronic pain in several of the UKBB body sites (face, head, neck/shoulder, back, stomach, pain all over the body) [80, 19]. The latter [19] examined the shared genetic overlap in pain (both chronic and short-term, i.e., less than a month) and neuropsychiatric disorders, which included several psychiatric conditions that we examined (bipolar disorder, schizophrenia, anorexia nervosa, anxiety, depression, PTSD, and ADHD). They found a similar pattern of genetic correlations to ours. Together with both of these studies, our findings show that the association between the incidence of different chronic pain conditions and high neuroticism may be at least in part due to their shared genetic risks, rather than a causal relationship between neuroticism and chronic pain. However, our finding is a correlational genetic relationship. To establish causality, additional studies are needed.

Our work goes beyond an examination of pairwise correlations between disorders and queries shared genetic variance as determined by formally extracted cross-condition factors. These factor-level genetic risk components are informative about the biological mechanisms shared among conditions that are not typically diagnosed or treated together or even by the same specialist (i.e. chronic pain by a primary care practitioner or neurologist, psychiatric disorders by a mental health specialist). Thus, the high correlation between chronic pain associations with neuroticism and inter-pain condition associations as well as the correlation between neuroticism and the General Pain factor also suggest that the biological mechanisms for greater susceptibility to cross-condition chronic pain may underlie an overall susceptibility to negative sensory and emotional experiences. The high inter-correlations observed between cross-condition chronic pain, externalizing and internalizing disorders, and neuroticism suggest that this susceptibility is characterized by the higher risks for substance abuse, depression, anxiety, PTSD, ADHD, as well as chronic pain.

Interestingly, the genetic risk common to cross-condition chronic pain and both the internalizing and externalizing condition factors is in large part attributable to all three categories of disorders sharing genetic risk with neuroticism. This overlap suggests that a potential avenue of intervention for people with internalizing and/or externalizing conditions as comorbidities is targeting symptoms of high neuroticism. Although the genetic correlations between chronic pain and both thought disorder factors are negligible, neuroticism appears to explain much of the relationship between chronic pain and the Psychotic factor. These results suggest that without high neuroticism, there is minimalto-no relationship between chronic pain and psychotic thought disorders.

There are several limitations in and attractive future directions that arise from our study. First, we provide a structurally informed model of genetic correlations among chronic pain, psychiatric conditions, and neuroticism. Although it is beyond the scope of this study, a functional annotation with relevant biological pathways will be an important next step to determine the specific mechanisms that underlie the genetic overlap between pain and psychiatric factors, as well as neuroticism. Second, it will be important to check the validity of our model by fitting it to data from independent samples. Third, the main populations used in our analyses are of White European descent, residing in Western Europe or the United States. It is our hope that as samples more representative of the world’s population continue to grow in size, we will be able to check how our model performs in people with other ancestral background and from other cultural settings.

In conclusion, the cross-condition overlapping genetic signals are informative about the risks for chronic pain onset that can be harnessed to prevent, treat, and mitigate suffering associated with pain. Our results underscore the importance of holistic clinical assessments that are informed by the presence of psychiatric problems, such as depression or anxiety (or even their symptoms, such as loneliness and miserableness) and by an assessment of the neuroticism personality dimension. Our results further support a conceptualization of chronic pain as closely related at the genetic level with psychiatric conditions, strengthening and expanding upon the biopsychosocial model. A joint treatment program that targets all comorbid conditions and traits with shared pathways of vulnerability may prove to be highly effective.

## Data Availability

All data produced in the present study are available upon reasonable request to the authors.

https://pgc.unc.edu/for-researchers/download-results/

## Notes

### Competing Interest Statement

The authors have declared no competing interest.

### Funding Statement

This study is funded by R01DA046064 and T32DA017637.

### Author Declarations

This study used publicly available summary association statistics from previously published genome-wide association studies.

